# Access, Affordability, and Quality of Medicines in Public Primary Health Facilities in Ghana: Implications for Rational Use of Medicines

**DOI:** 10.64898/2026.05.14.26353169

**Authors:** Dziedzom Kwesi Awalime, Genevieve Cecilia Aryeetey, Augustina Koduah

**Author notes:** **Corresponding author:** Dziedzom Kwesi Awalime, Ghana College of Physicians and Surgeons, Department of Public Health, 1 Jomo Kenyatta Road, Ridge. P. O. Box MB 429, Accra. **Note on corresponding author affiliation:** The corresponding author conducted this research while enrolled in the Master of Health Economics program at the University of Ghana. The affiliation above reflects the author’s position at the time of submission.

## Abstract

Rational use of medicines (RUM) is a global health priority, yet significant challenges persist in low- and middle-income countries (LMICs), particularly around medicine access, affordability, and quality. While RUM studies often focus on prescribing practices, systemic barriers such as supply chain inefficiencies and pricing receive less attention. This study assessed three key health system components of RUM (availability, affordability, and quality of essential medicines) at two public primary health facilities in Ghana and examined patient care practices against WHO RUM standards. A quantitative, cross-sectional study was conducted at Kekele Polyclinic and Rawlings Circle Polyclinic in Accra. Retrospective data were extracted from prescription sheets, medicine tally cards, and ledgers to evaluate WHO Level II core drug use indicators. Fifteen essential medicines were selected based on the Ghana Essential Medicines List, Standard Treatment Guidelines, and municipal disease burden data. Exit interviews with 107 patients assessed dispensing and counselling practices, and structured observation covered storage conditions and pharmaceutical handling. Availability of key medicines fell significantly short of WHO targets, with Rawlings Circle meeting only 40% and Kekele 73.3% of the 100% benchmark. Treatment of malaria and pneumonia cost patients up to three times the national daily minimum wage, indicating poor affordability. The average number of medicines prescribed per encounter (3.2) exceeded the WHO recommended standard (≤2). Storage and handling infrastructure was inadequate, with both facilities falling short of recommended conservation standards. Gaps in medicine availability, affordability, and infrastructure undermine rational medicine use in primary healthcare. Strengthening procurement systems, enforcing storage protocols, and implementing financial protection mechanisms are essential for equitable and safe medicine use within Ghana’s health system.

## Introduction

Access to essential medicines is fundamental to effective healthcare delivery and is integral to achieving the rational use of medicines (RUM). According to the World Health Organization (WHO), RUM is achieved when “patients receive medications appropriate to their clinical needs, in doses that meet their own individual requirements, for an adequate period, and at the lowest cost to them and the community.” ^[1]^ Despite being a critical component of universal health coverage, rational medicine use is still inadequately implemented across many LMICs, where medicine-related barriers disproportionately affect population health.

Globally, over half of all medicines are prescribed or dispensed inappropriately, and nearly 50% of patients fail to take their medications as intended. ^[2]^ Contributing factors include polypharmacy, overuse of injections, non-compliance with clinical guidelines, and self-medication. ^[3,4]^ Equally important but often overlooked are systemic issues such as medicine unavailability, inadequate storage infrastructure, and high out-of-pocket (OOP) costs of treatment, all of which can exacerbate irrational use. In sub-Saharan Africa, as much as 90% of the population purchases medicines through OOP payments, making pharmaceuticals the second-largest household expense after food. ^[5]^ Global evidence demonstrates that public sector availability of essential medicines averages as low as 35% across LMICs. ^[6]^

In Ghana, primary healthcare (PHC) facilities play a frontline role in ensuring equitable access to essential health services. Persistent challenges including frequent stockouts, affordability gaps, and suboptimal medicine storage undermine the capacity of these facilities to support rational medicine use. Although several studies in Ghana have evaluated prescribing trends, ^[7,8]^ few have comprehensively examined the combined effect of access, affordability, and conservation conditions on RUM. A health systems perspective, which situates medicine access within the broader functions of financing, service delivery, and supply chain governance, is essential for identifying and addressing such structural gaps. ^[9,10]^

This study addresses that gap by evaluating the core health system components influencing rational medicine use in two public PHC facilities in the Madina municipality. Specifically, it aims to:

1. Assess the availability and affordability of essential medicines for treating common conditions including malaria, pneumonia, hypertension, and diabetes.
2. Examine medicine storage and conservation infrastructure as a proxy for medicine quality.
3. Evaluate patient care practices including prescribing patterns, medicine labelling, and patient knowledge in alignment with WHO core indicators for RUM.

By adopting a systems-oriented approach, this study contributes new insights into the operational challenges affecting rational medicine use in urban PHC settings in Ghana.

## Materials and Methods

### Study Design and Setting

This was a descriptive, cross-sectional study using quantitative methods to evaluate rational medicine use in two public primary health facilities (Kekele Polyclinic and Rawlings Circle Polyclinic) in the La Nkwantanang-Madina Municipality of Greater Accra, Ghana. The two facilities are situated within 1.5 km of each other in a busy urban district adjacent to one of Accra’s largest trading hubs. Both facilities serve a diverse urban population with a high burden of infectious and chronic diseases. This study is reported in accordance with the Strengthening the Reporting of Observational Studies in Epidemiology (STROBE) guidelines for cross-sectional studies ^[11]^; a completed STROBE checklist is provided as S1 Checklist.

### Study Population and Sampling

The study population comprised outpatient clients (n = 107), clinicians (n = 2), dispensers (n = 2), pharmacists (n = 4), and pharmacy store managers (n = 2) present during the survey period. Clients were eligible if they had completed a consultation and obtained prescribed medicines from the outpatient pharmacy. Inclusion was restricted to general outpatient cases such as malaria, pneumonia, hypertension, and diabetes. Specialised services (e.g., antenatal/postnatal care, child welfare clinics, and specialist referrals) were excluded due to data interpretation challenges.

Systematic random sampling was used to select clients exiting the pharmacy. Each was interviewed using a structured tool adapted from WHO’s Level II Facility Survey on drug use indicators.

### Data Collection Tools and Indicators

WHO’s core drug use indicators were used to assess access, affordability, quality, and patient care practices. The full list of indicators and their assessments is presented in Table 1, and the WHO optimal values are listed in Table 2.

**Table 1:**
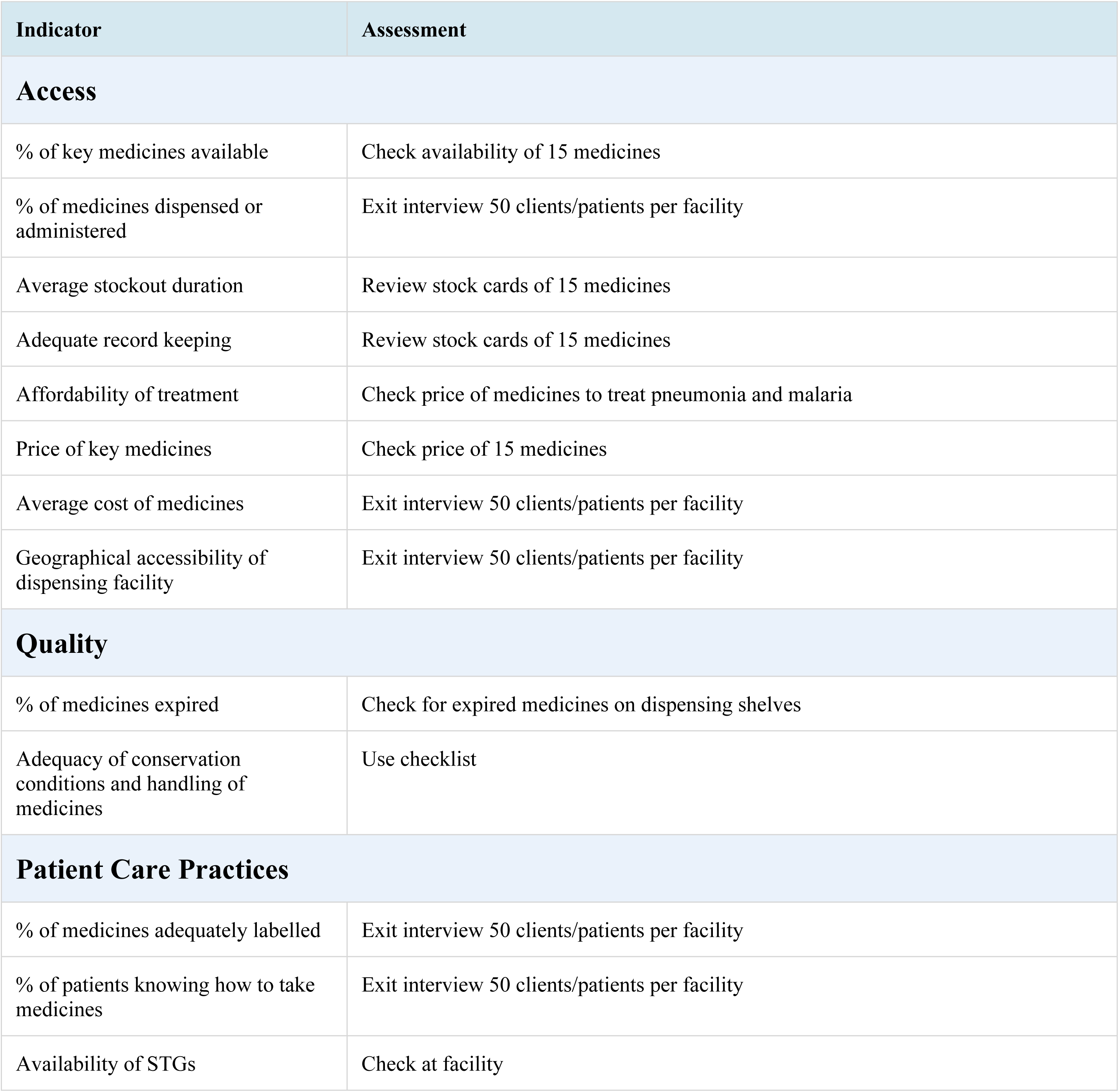
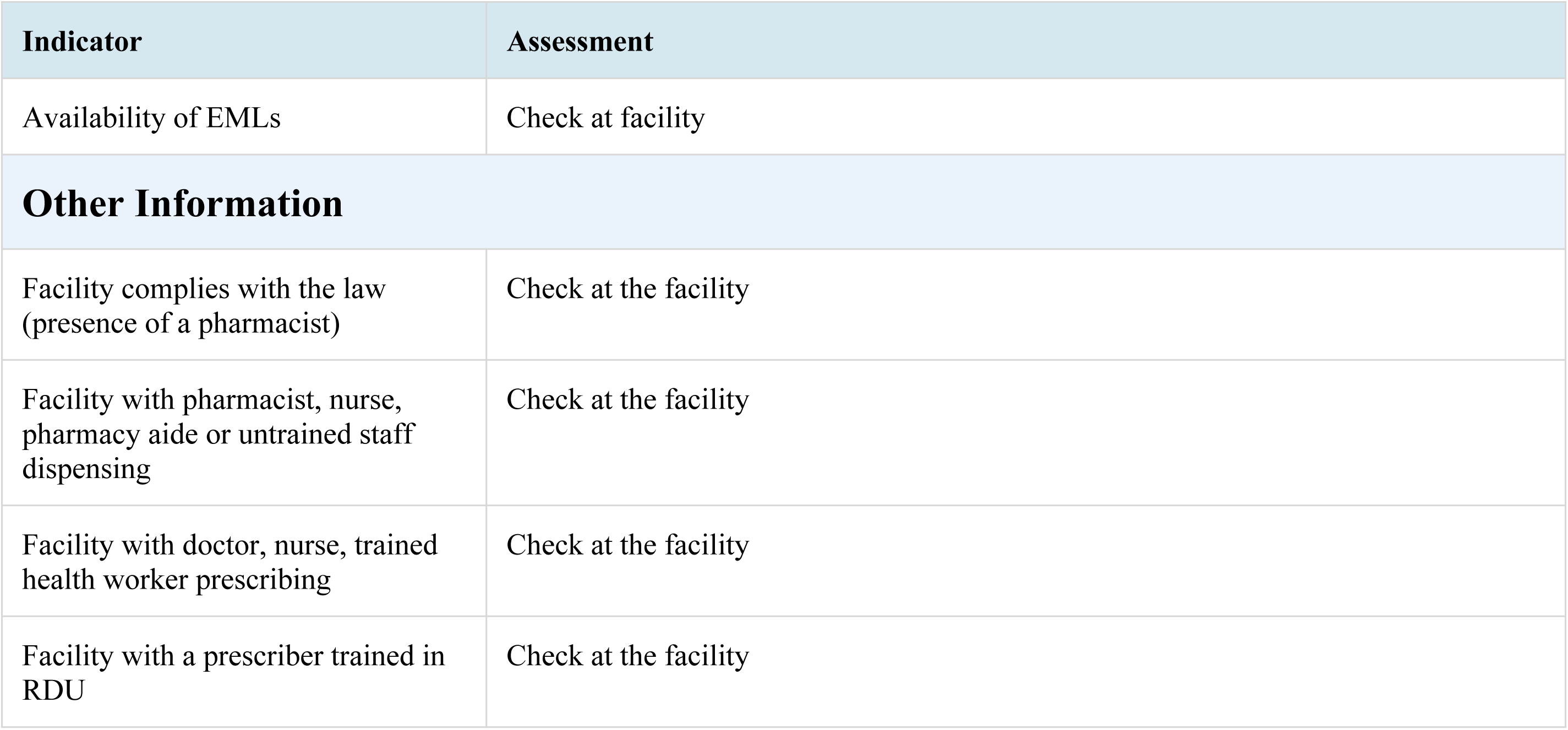
Indicator list for survey.

| Indicator | Assessment |
| --- | --- |
| **Access** |  |
| % of key medicines available | Check availability of 15 medicines |
| % of medicines dispensed or administered | Exit interview 50 clients/patients per facility |
| Average stockout duration | Review stock cards of 15 medicines |
| Adequate record keeping | Review stock cards of 15 medicines |
| Affordability of treatment | Check price of medicines to treat pneumonia and malaria |
| Price of key medicines | Check price of 15 medicines |
| Average cost of medicines | Exit interview 50 clients/patients per facility |
| Geographical accessibility of dispensing facility | Exit interview 50 clients/patients per facility |
| **Quality** |  |
| % of medicines expired | Check for expired medicines on dispensing shelves |
| Adequacy of conservation conditions and handling of medicines | Use checklist |
| **Patient Care Practices** |  |
| % of medicines adequately labelled | Exit interview 50 clients/patients per facility |
| % of patients knowing how to take medicines | Exit interview 50 clients/patients per facility |
| Availability of STGs | Check at facility |
| Availability of EMLs | Check at facility |
| **Other Information** |  |
| Facility complies with the law (presence of a pharmacist) | Check at the facility |
| Facility with pharmacist, nurse, pharmacy aide or untrained staff dispensing | Check at the facility |
| Facility with doctor, nurse, trained health worker prescribing | Check at the facility |
| Facility with a prescriber trained in RDU | Check at the facility |

**Table 2:** Core medicine use indicators and their WHO optimal values.

| Core Medicine Use Indicators | WHO Optimal Values |
| --- | --- |
| **Prescribing Indicators** |  |
| Average number of medicines prescribed per patient encounter | 1.6–1.8 |
| **Patient-Care Indicators** |  |
| Percent medicines dispensed | 100% |
| Percent medicines adequately labelled | 100% |
| Percent patients with knowledge of correct doses | 100% |
| **Facility-Specific Indicators** |  |
| Availability of essential medicines list or formulary to practitioners | 100% |
| Percent key medicines available | 100% |

Fifteen key medicines were selected in consultation with the Municipal Health Directorate based on the top five prevalent conditions in the area and cross-verified with the 2017 Ghana Essential Medicines List (EML) and Standard Treatment Guidelines (STG).

### Data Collection Procedure

Data collection was carried out over a one-week period between 25^th^ to 29^th^ September 2023. Prescription data, stock records, and medicine tally cards were reviewed retrospectively for the prior 12 months (between August 2022 and August 2023). Exit interviews were conducted with clients using structured digital forms uploaded to CommCare on mobile tablets. All participants were given a written informed consent form which they signed. No patient identifiers like names, hospital number or insurance number were collected, only demographic information like age, sex etc. were collected. Authors did not require patient identifier information for analysis.

### Data Management and Analysis

Quantitative data were cleaned and analysed using Microsoft Excel and STATA version 16. Descriptive statistics (means, percentages, and frequencies) were calculated. Medicine costs were converted to USD using the average interbank exchange rate for June 2023 (GHS 11.06 = USD 1). Affordability was estimated by comparing treatment costs with the national daily minimum wage (GHS 14.88 in 2023).

### Ethical Considerations

Ethical approval was obtained from the Ghana Health Service Ethics Review Committee (GHS-ERC: 025/06/23). Administrative clearance was granted by the Greater Accra Regional Health Directorate and the La Nkwantanang-Madina Municipal Health Directorate. Written informed consent was obtained from all participants. Exit interviews were conducted in English or local languages (Twi, Ga, or Ewe) depending on patient preference. No personal identifiers were recorded, and all data were anonymized to ensure confidentiality. This study was conducted in accordance with the Declaration of Helsinki.

## Results

### Participant and Facility Characteristics

A total of 107 outpatients were interviewed: 51 at Madina Polyclinic – Rawlings Circle and 56 at Madina Polyclinic – Kekele. Women represented 74.8% of respondents, and mean ages were 36 years (Rawlings Circle) and 33 years (Kekele). Young adults (aged 19–35 years) formed the largest age category (42.1%). Both facilities had pharmacists on-site and were compliant with national legal requirements for staffing and reference materials, including the Ghana EML and STG. The sex and age distributions of respondents are presented in Tables 3 and 4.

**Table 3:** Tabulation of respondent sex by facility.

| Facility | Sex | Frequency | Percent (%) |
| --- | --- | --- | --- |
| Both Facilities | Female | 80 | 74.77 |
|  | Male | 27 | 25.23 |
|  | Total | 107 | 100.00 |
| Rawlings Circle | Female | 41 | 80.39 |
|  | Male | 10 | 19.61 |
|  | Total | 51 | 100.00 |
| Kekele | Female | 39 | 69.64 |
|  | Male | 17 | 30.36 |
|  | Total | 56 | 100.00 |

**Table 4:** Tabulation of respondent age by facility.

| Facility | Age Category | Frequency | Percent (%) |
| --- | --- | --- | --- |
| Both Facilities | Children | 19 | 17.76 |
|  | Young Adults | 45 | 42.06 |
|  | Older Adults | 35 | 32.71 |
|  | Aged | 8 | 7.48 |
|  | Total | 107 | 100.00 |
| Rawlings Circle | Children | 3 | 5.88 |
|  | Young Adults | 26 | 50.98 |
|  | Older Adults | 19 | 37.25 |
|  | Aged | 3 | 5.88 |
|  | Total | 51 | 100.00 |
| Kekele | Children | 16 | 28.57 |
|  | Young Adults | 19 | 33.93 |
|  | Older Adults | 16 | 28.57 |
|  | Aged | 5 | 8.93 |
|  | Total | 56 | 100.00 |

### Access to Essential Medicines

#### Availability of Key Medicines

Availability of the 15 key tracer medicines was 73.3% at Kekele and 40% at Rawlings Circle; both fall short of the WHO 100% optimal benchmark. Only six tracer medicines were available in both facilities at the time of the survey. Magnesium trisilicate, ferrous sulphate, artesunate-amodiaquine, and omeprazole were absent from both.

#### Stockouts

Stockout durations ranged from 0 to 240 days. In Rawlings Circle, three medicines were consistently in stock throughout the prior year, while in Kekele only cefuroxime had no recorded stockout. Seven of 15 tracer medicines at Kekele were unavailable for over 100 days; the most affected were ferrous sulphate, oral rehydration salts, and omeprazole.

### Geographic Accessibility

Most clients (94.3%) reached the facilities within an hour; 55% of Rawlings Circle users arrived in under 30 minutes, while 65% of Kekele clients arrived within 30–60 minutes. Transport cost averaged GHS 10.32 (USD 0.93) to Rawlings Circle and GHS 8.88 (USD 0.80) to Kekele, representing 69.3% and 59.7% of the daily minimum wage, respectively. Travel time data are presented in Table 5.

**Table 5:** Tabulation of time to reach the facility.

| Travel Time | Rawlings Circle | Kekele | Total |
| --- | --- | --- | --- |
| Less than 30 minutes | 28 (55%) | 19 (37%) | 47 |
| 30 minutes to 1 hour | 21 (41%) | 33 (65%) | 54 |
| More than 1 hour | 2 (4%) | 4 (8%) | 6 |
| Total | 51 | 56 | 107 |

### Affordability of Treatment

Cost analysis of treatment for pneumonia and malaria revealed substantial affordability gaps. Full treatment cost data are provided in Tables 6 and 7; summary findings are as follows.

**Table 6:** Cost of treatment for common conditions (full data in S2 Appendix) Note: Full cost breakdown is provided in S2 Appendix. Daily minimum wage = GHS 14.88. 2023 interbank exchange rate: GHS/USD = 11.06.

**Table 7:** Cost of essential medicines at Kekele and Rawlings Circle (full data in S2 Appendix) *Note: Full medicine price table is provided in S2 Appendix*.

Pneumonia (adult): Rawlings Circle: GHS 44.68 (USD 4.04; approximately 3 days’ wages); Kekele: GHS 37.40 (USD 3.38; approximately 2.5 days’ wages).

Pneumonia (child under 5): Rawlings Circle: GHS 17.00 (USD 1.54; approximately 1.1 days’ wage); Kekele: GHS 25.00 (USD 2.26; approximately 1.7 days’ wages).

Malaria (adult): Rawlings Circle: GHS 5.52 (USD 0.50; 0.37 days’ wage); Kekele: GHS 3.78 (USD 0.34; 0.25 days’ wage).

Malaria (child under 5): Rawlings Circle: GHS 34.02 (USD 3.08; 2.3 days’ wages); Kekele: GHS 34.98 (USD 3.16; 2.4 days’ wages).

All treatment costs exceeded WHO’s affordability threshold of one day’s wage.

### Quality of Medicines and Storage Conditions

No expired medicines were found on dispensing shelves at either facility. Record-keeping for key medicines was complete for 86.7% of tracer drugs in Rawlings Circle and 80% in Kekele.

In Rawlings Circle, the dispensing area met all storage adequacy criteria, but the storeroom fell short due to moisture from ceiling leakage. In Kekele, cold storage lacked regular temperature logging. All other storage indicators, first-expiry-first-out (FEFO) rotation, systematic organization, and pest control were met at both facilities. Full conservation data are presented in Table 8.

**Table 8:** Adequacy of conservation conditions at Rawlings Circle and Kekele.

| Storage Assessment Indicator | RC Storeroom | RC Dispensary | KK Storeroom | KK Dispensary |
| --- | --- | --- | --- | --- |
| Temperature control method in place | 1 | 1 | 1 | 1 |
| Windows or air vents present | 1 | 1 | 1 | 1 |
| Direct sunlight blocked | 1 | 1 | 1 | 1 |
| Area free from moisture | 0 | 1 | 1 | 1 |
| Cold storage available | 1 | 1 | 1 | 1 |
| Regularly filled temperature chart | 1 | 1 | 0 | 0 |
| Medicines not stored on floor | 1 | – | 1 | – |
| Medicines stored systematically | 1 | 1 | 1 | 1 |
| Medicines stored FEFO | 1 | 1 | 1 | 1 |
| No evidence of pests | 1 | 1 | 1 | 1 |
| Tablets not handled by naked hand | – | 1 | – | 1 |
| *Note: 1 = adequate; 0 = inadequate; – = not applicable. RC = Rawlings Circle; KK = Kekele.* |  |  |  |  |

### Patient Care Practices

Descriptive statistics for all patient-level variables are presented in Table 9.

**Table 9:** Descriptive statistics by facility.

| Variable | Facility | Obs. | Mean | Std. Dev. | Min | Max |
| --- | --- | --- | --- | --- | --- | --- |
| Age (years) | Both | 107 | 35.12 | 18.50 | 1 | 89 |
|  | RC | 51 | 36.52 | 15.29 | 3 | 70 |
|  | KK | 56 | 33.84 | 21.06 | 1 | 89 |
| # Medicines Prescribed | Both | 107 | 3.22 | 1.41 | 1 | 8 |
|  | RC | 51 | 3.45 | 1.71 | 1 | 8 |
|  | KK | 56 | 3.02 | 1.04 | 1 | 5 |
| # Medicines Dispensed | Both | 107 | 2.22 | 1.17 | 0 | 6 |
|  | RC | 51 | 2.27 | 1.28 | 0 | 6 |
|  | KK | 56 | 2.16 | 1.08 | 0 | 5 |
| # Adequately Labelled | Both | 107 | 2.16 | 1.19 | 0 | 6 |
|  | RC | 51 | 2.21 | 1.30 | 0 | 6 |
|  | KK | 56 | 2.11 | 1.09 | 0 | 5 |
| Amount Paid (GHS) | Both | 107 | 47.08 | 55.23 | 0 | 284 |
|  | RC | 51 | 59.31 | 68.33 | 0 | 284 |
|  | KK | 56 | 35.95 | 37.01 | 0 | 120 |
| Transport Cost (GHS) | Both | 107 | 9.57 | 12.77 | 0 | 100 |
|  | RC | 51 | 10.32 | 11.32 | 0 | 50 |
|  | KK | 56 | 8.88 | 14.03 | 0 | 100 |
| *RC = Rawlings Circle; KK = Kekele.* |  |  |  |  |  |  |

Prescribing trends showed an average of 3.2 medicines per encounter (Rawlings Circle: 3.4; Kekele: 3.02), both exceeding the WHO recommended threshold of ≤2, suggesting potential polypharmacy. Dispensing rates were 68.7% below the WHO optimal of 100% and below national benchmarks. A total of 97.5% of medicines were adequately labelled. Of the 107 patients, 91.6% could correctly describe how to take their medicines (Rawlings Circle: 98.1%; Kekele: 85.7%). Patient knowledge data by facility and sex are presented in Tables 10 and 11.

**Table 10:** Patient/caregiver knowledge of medicine use by facility.

| Facility | Knows How to Take Medicines | Frequency | Percent (%) |
| --- | --- | --- | --- |
| Both Facilities | No | 9 | 8.41 |
|  | Yes | 98 | 91.59 |
|  | Total | 107 | 100.00 |
| Rawlings Circle | No | 1 | 1.96 |
|  | Yes | 50 | 98.04 |
|  | Total | 51 | 100.00 |
| Kekele | No | 8 | 14.29 |
|  | Yes | 48 | 85.71 |
|  | Total | 56 | 100.00 |

**Table 11:**
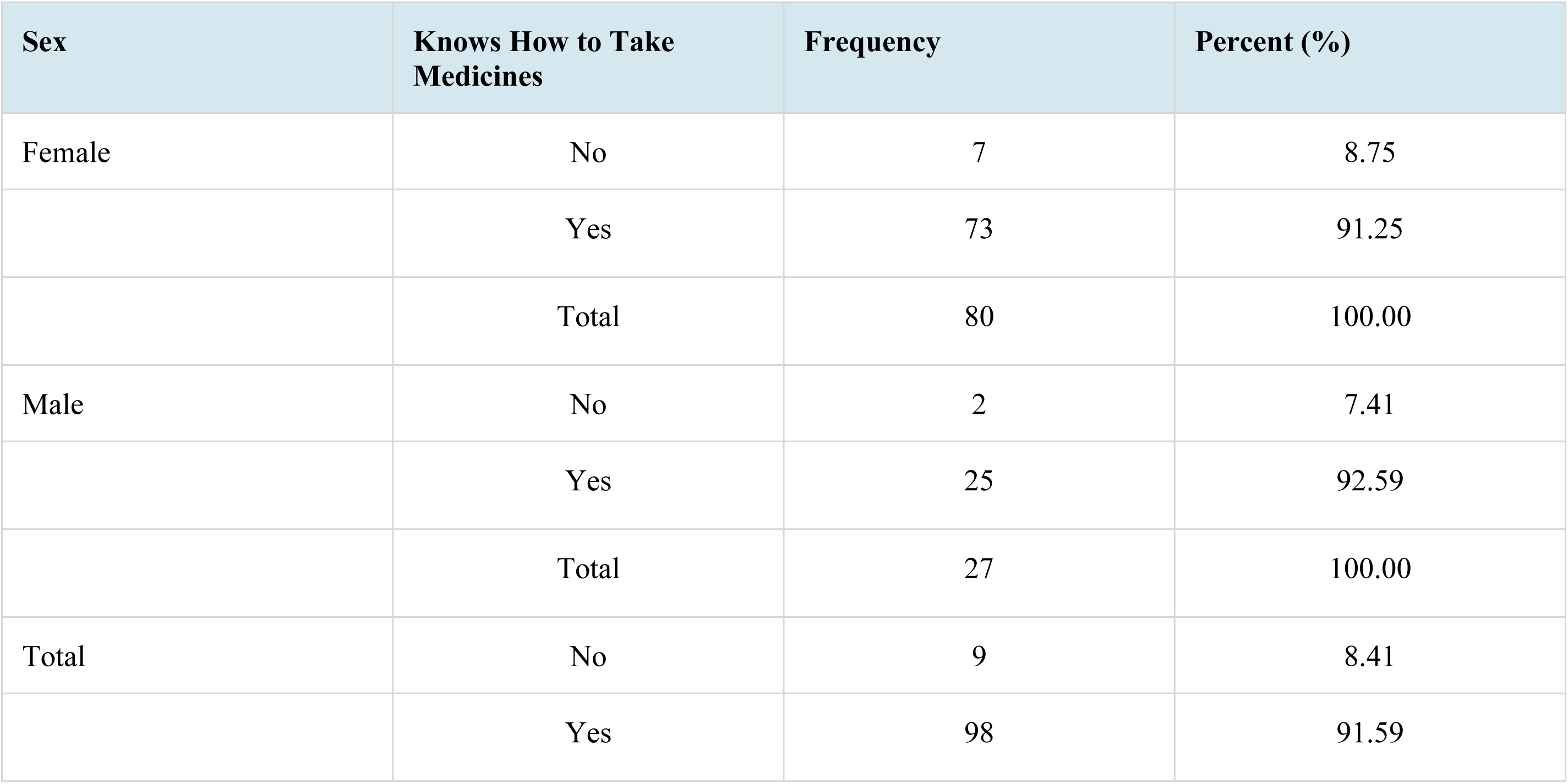
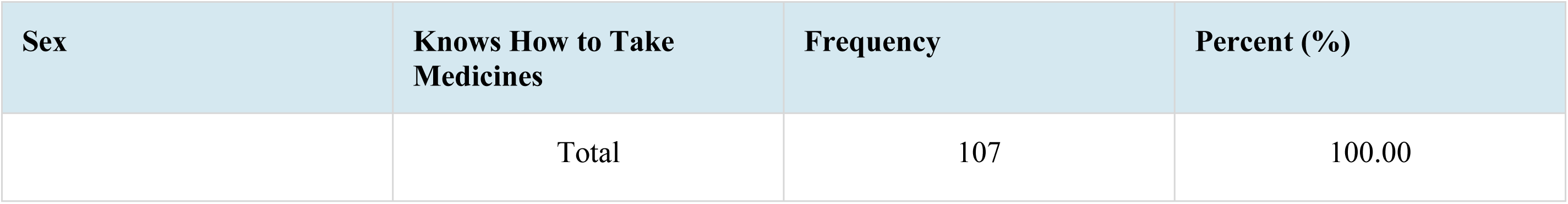
Patient knowledge of medicine use by sex.

| Sex | Knows How to Take Medicines | Frequency | Percent (%) |
| --- | --- | --- | --- |
| Female | No | 7 | 8.75 |
|  | Yes | 73 | 91.25 |
|  | Total | 80 | 100.00 |
| Male | No | 2 | 7.41 |
|  | Yes | 25 | 92.59 |
|  | Total | 27 | 100.00 |
| Total | No | 9 | 8.41 |
|  | Yes | 98 | 91.59 |
|  | Total | 107 | 100.00 |

## Discussion

This study assessed rational medicine use from the perspective of access, affordability, and quality in two urban primary healthcare facilities in Ghana. While previous studies have emphasized inappropriate prescribing and self-medication practices, ^[3,4]^ our results show that systemic barriers including medicine unavailability, conservation infrastructure gaps, and high treatment costs also play a decisive role in RUM outcomes at the facility level.

### Access to Medicines

Medicine availability was below the WHO 100% benchmark at both sites, with Rawlings Circle meeting only 40% and Kekele 73.3% of the tracer medicine criteria. These findings mirror evidence from similar contexts in Ghana and other LMICs, where stockouts and erratic supply chains compromise care quality. ^[12,14]^ Compared with the median national stockout duration of 29.9 days reported in Ghana’s 2012 pharmaceutical sector assessment, the average stockout of 136 days at Kekele is alarmingly high. This is consistent with the global pattern identified by Cameron et al., who found public sector tracer medicine availability averaging below 40% across 36 LMICs. ^[6]^

Poor availability erodes patient trust in public facilities and shifts demand toward private vendors, often at higher cost and with questionable medicine quality. ^[15]^ Public sector stockouts also drive-up private sector prices, creating a compounding affordability burden. ^[13]^ These findings underscore the need for improved procurement systems, supply forecasting, and inventory management.

### Affordability of Treatment

Affordability remains a critical barrier to accessing medicines in systems dominated by OOP payments. Treatment for adult pneumonia cost up to three times the daily minimum wage, a burden that can delay care or push households into poverty, as documented in Ethiopia and Uganda. ^[5,16]^ WHO recommends that essential treatments should not exceed a single day’s wage, a threshold exceeded by nearly all treatments in this study, particularly for children’s malaria.

Although geographic access was generally good (94.3% of patients arrived within one hour), transportation costs consumed up to 69% of the daily wage. Evidence from Nigeria and Chad documents similar patterns, where transport costs significantly limit health-seeking behaviour among the urban poor. ^[17,18]^ The COVID-19 pandemic further highlighted how transport disruptions widen access inequities. ^[19]^

### Quality of Medicines and Infrastructure

Despite no expired drugs on pharmacy shelves, important storage gaps were identified. Moisture from ceiling leakage at Rawlings Circle and irregular cold-chain temperature logging at Kekele create risks for medicine stability. Inadequate storage can reduce the therapeutic value of medicines or increase adverse effects. ^[20,21]^ Neither facility achieved WHO’s 100% conservation standard, though both exceeded national averages from Ghana’s 2012 pharmaceutical sector survey.

### Patient Care Practices

The average of 3.2 medicines per encounter exceeds WHO’s threshold (≤2), indicating a polypharmacy trend that increases the risk of adverse drug events and non-adherence. ^[22,23]^ A systematic review across the WHO African region found averages of 2.6–3.1 medicines per encounter, placing the current findings at the upper end of the regional range. ^[14]^ Although lower than Sierra Leone ^[24]^ and Northern Ghana, ^[25]^ the finding still warrants attention through prescriber training and reinforced STG use.

Dispensing rates (68.7%) were below national (80%) and international benchmarks, Egypt reports 95.9% ^[26]^ likely driven by stockouts. Patient knowledge of medicine use was, however, high (91.6%), indicating effective pharmacist communication even under resource constraints.

### Policy Implications

The findings point to four priority areas for strengthening Ghana’s primary healthcare pharmaceutical services:

4. Improved procurement and stock management systems to reduce stockout durations;
5. Targeted subsidies for essential medicines to reduce OOP burden, particularly for childhood malaria and pneumonia;
6. Capital investment in pharmacy infrastructure, especially cold chain maintenance and storeroom repair;
7. Continuous professional development for prescribers and dispensers on RUM principles and STG adherence.

Integrating these priorities into Ghana’s national pharmaceutical policy will be critical for achieving equitable, efficient, and safe medicine use in line with Universal Health Coverage goals.

## Conclusion

This study documents persistent challenges in achieving rational medicine use at the primary healthcare level in Ghana. Both facilities showed varying degrees of alignment with WHO core RUM standards, with notable gaps in medicine availability, affordability, and storage infrastructure.

Availability of key medicines fell substantially below WHO benchmarks, stockout durations were prolonged, and treatment costs frequently exceeded WHO’s affordability threshold. Patient knowledge of medicine use and labelling compliance were encouraging, but high prescribing volumes and conservation shortfalls signal systemic weaknesses that risk undoing good dispensing practice.

Addressing these weaknesses requires coordinated action across supply chain management, pharmaceutical financing, and healthcare infrastructure, reforms that would bring Ghana closer to its Universal Health Coverage targets and provide a replicable model for similar urban PHC settings across sub-Saharan Africa.

## Study Limitations

This study has several limitations. First, the accuracy of stockout durations may have been affected by incomplete or inconsistent facility record-keeping. Second, patient-reported travel time may be biased by the proximity of a major market; many clients may have departed from the market rather than from home, potentially overestimating geographic accessibility. Third, local medicine prices were not compared to international reference prices, as the MSH Drug Price Indicator Guide has not been updated since 2015. Finally, the study was conducted in only two urban facilities, limiting generalizability to rural or peri-urban settings.

Despite these limitations, the study provides actionable, facility-level evidence on systemic barriers to rational medicine use and offers a replicable assessment framework for similar settings.

## Data availability

All data used in the paper has been presented in the manuscript. Any additional data required shall be provided upon reasonable request.

## Acknowledgments

The authors thank the management and staff of Madina Polyclinic – Rawlings Circle and Madina Polyclinic – Kekele for their cooperation and support during data collection. The La Nkwantanang-Madina Municipal Health Directorate is acknowledged for providing municipal health data and facilitating access to the study sites. The field assistants who administered the exit interviews are thanked for their diligence and professionalism.

---

**Fig 1.**
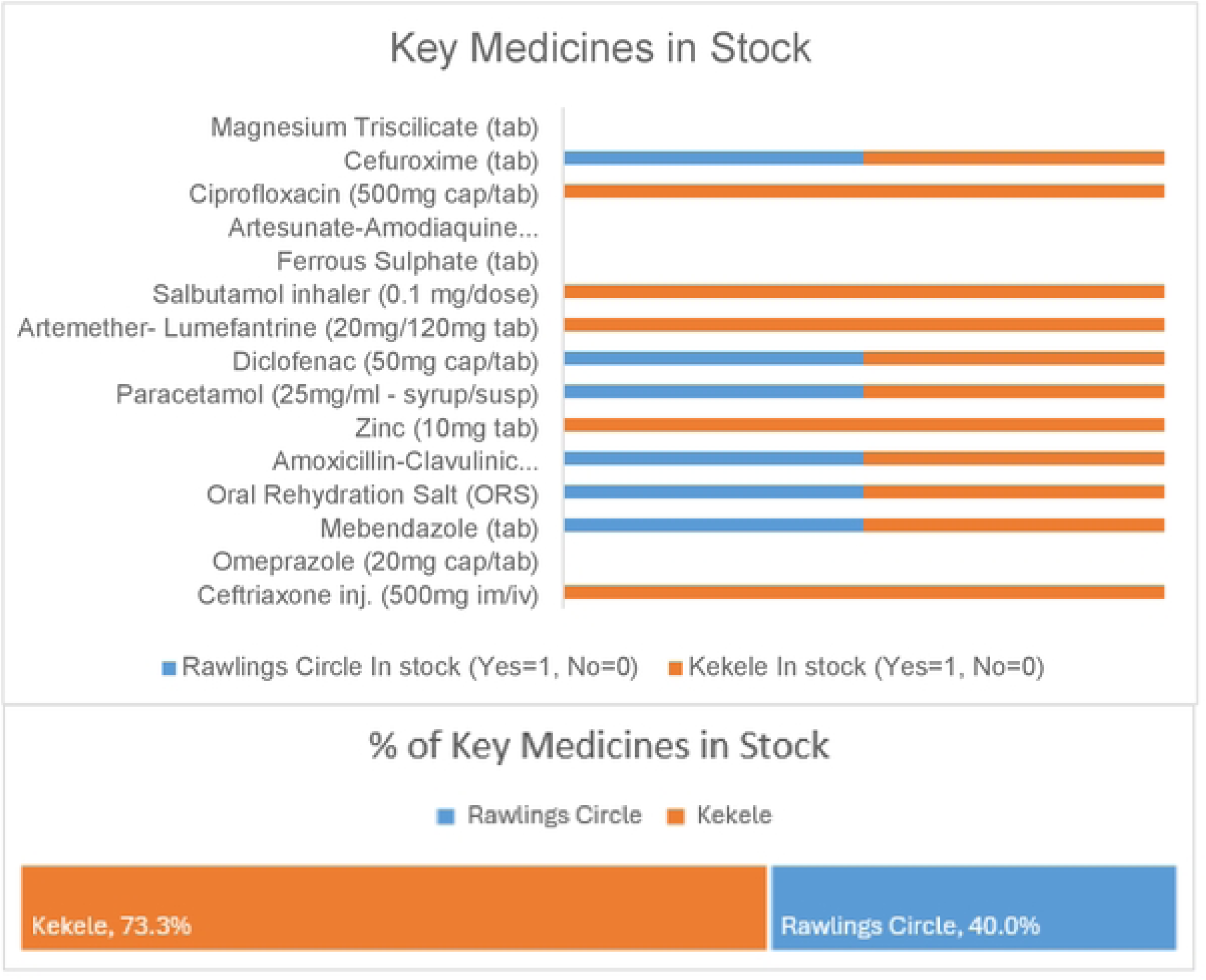
Stock of key medicines at both facilities

**Fig 2.**
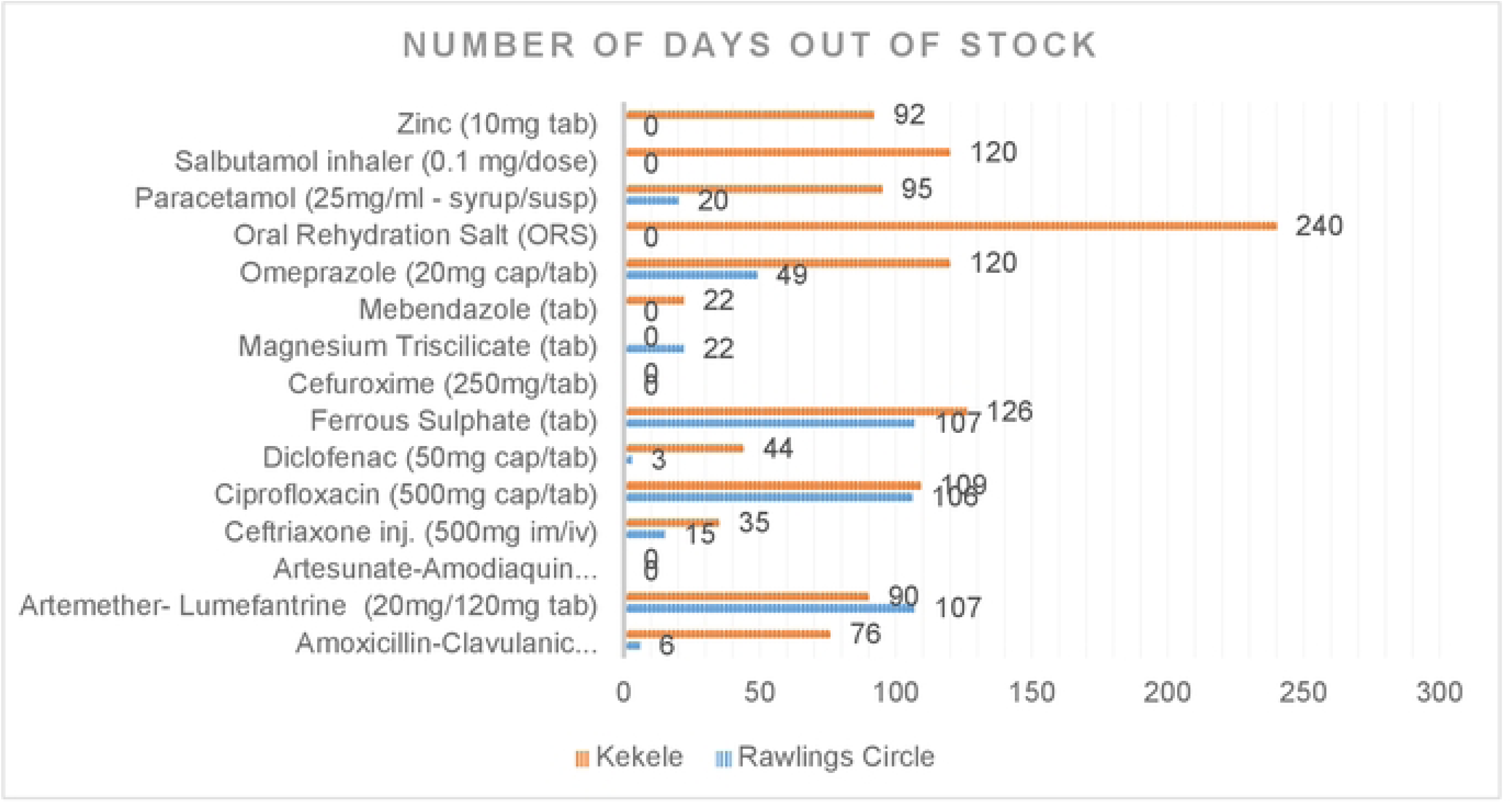
Stockout duration of key medicines at both facilities

**Fig 3.**
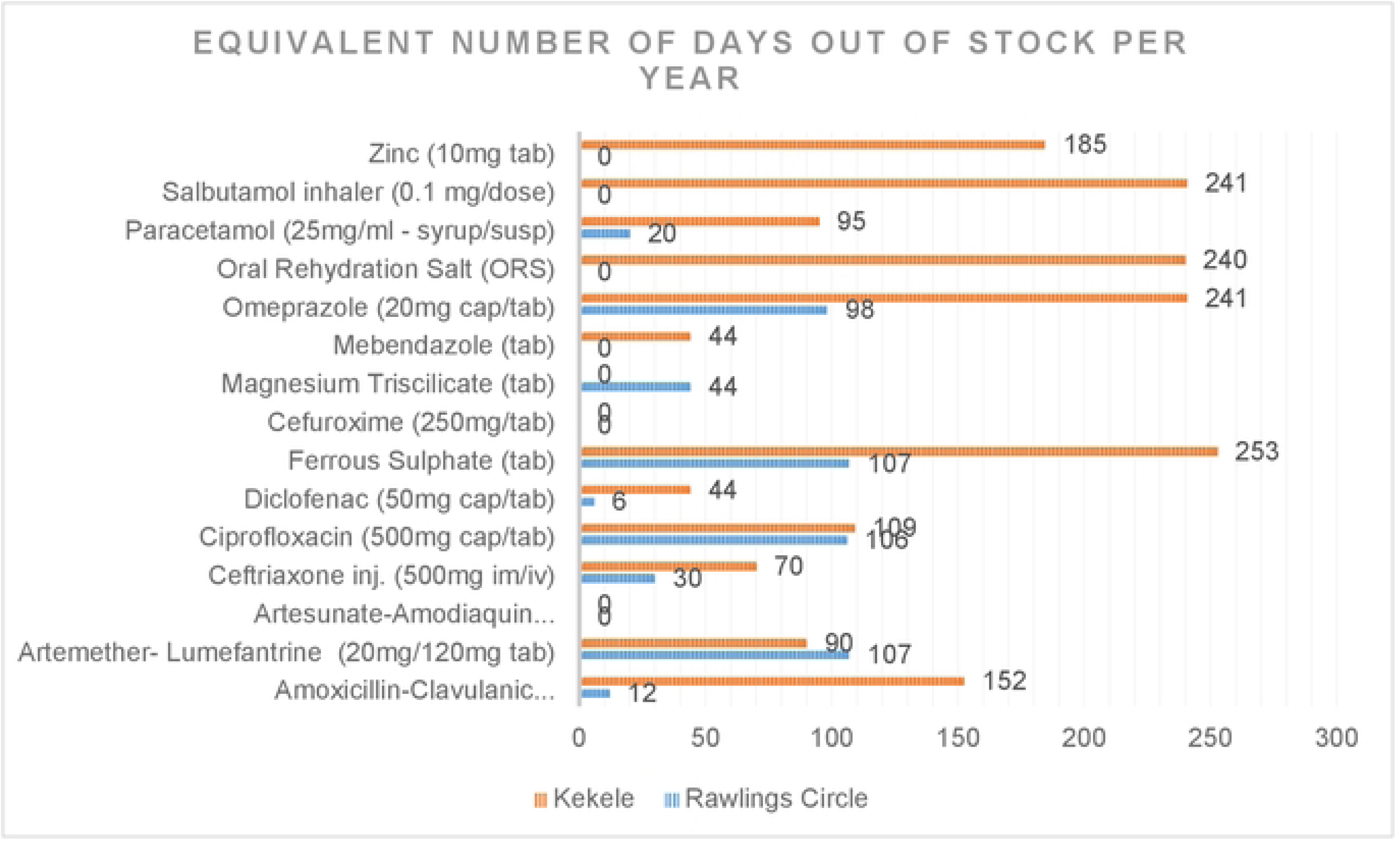
Equivalent yearly stockout period

## Notes

### Competing Interest Statement

The authors have declared no competing interest.

### Funding Statement

The research was self-funded as part of an academic requirement for completion of a Master of Health Economics program at the University of Ghana.

### Author Declarations

Ghana Health Service Ethics Review Committee GHS-ERC NUMBER: GHS-ERC: 025/06/23

